# Real-world effectiveness and non-inferiority evaluation and comparison of mRNA- and protein-based COVID-19 vaccines: a randomized study protocol (BEEHIVE)

**DOI:** 10.1101/2025.07.21.25331726

**Authors:** Sarang K. Yoon, German L. Ellsworth, Steph Battan-Wraith, Andrew L. Phillips, Rebecca V. Fink, Joshua Griffin, Elizabeth A.K. Rowley, Jacob McKell, Ashley S. Smith, Riley Campbell, Jesse Williams, Sarah W. Ball, Hongwei Zhao, Brandy Warren, Matthew D. Rousculp, Matthew S. Thiese

**Author notes:** **Corresponding author** Sarang K. Yoon, DO, MOH, University of Utah School of Medicine, Rocky Mountain Center for Occupational and Environmental Health, 250 E 200 S Suite 100, Salt Lake City, Utah 84111.

## Abstract

**Background:** Surveillance of COVID-19 vaccine effectiveness was extensive upon vaccine introduction; however, it slowed once pandemic status was withdrawn in May 2023. Continued monitoring of updated vaccine formulations is needed to ensure maintenance of vaccine effectiveness (VE) in the face of evolving viral strains.

**Objective:** The randomized, controlled BEEHIVE study (NCT06065176) was developed to assess real-world VE of the 2023–2024 Pfizer-BioNTech and Novavax COVID-19 vaccine formulations, which are directed to the XBB.1.5 SARS-CoV-2 variant.

**Methods:** This study was designed to enroll ∼1500 participants aged ≥18 years from the Salt Lake City, Utah area who had previously received ≥2 doses of an authorized mRNA-based COVID-19 vaccine, but not a dose of the 2023–2024 formulation. Study groups included two blinded groups randomized to receive the 2023–2024 formula of either the Novavax COVID-19 Vaccine or Pfizer-BioNTech COVID-19 Vaccine and a non-randomized comparator group of volunteers who chose not to receive a 2023–2024 vaccine dose during the study. Follow-up lasted 24 weeks and included symptom surveys and self-administered COVID-19 antigen testing, both occurring weekly. The primary aim was to compare VE (preventing symptomatic SARS-CoV-2 infection) between study-vaccinated participants and the comparator group. The secondary aim was to determine the relative VE of the Pfizer-BioNTech mRNA and Novavax 2023–2024 COVID-19 vaccines. Secondary objectives included assessment of how the number of prior COVID-19 vaccinations impacted VE of the 2023–2024 COVID-19 vaccines, predictors and associated factors for asymptomatic versus symptomatic infection and/or prolonged or severe illness, factors associated with post-COVID conditions (PCC), and knowledge, attitudes, and practices of participants related to COVID-19 vaccination. Participant engagement was maintained via online and text-based reminders and surveys, and researcher follow-up.

**Results:** Participants were recruited from November 2023 through March 2024 with 452 and 457 randomized to the Novavax and Pfizer-BioNTech vaccine groups, respectively, and 279 enrolled into the comparator group. SARS-CoV-2 variants from the XBB, JN.1, KP.2, and KP.3 lineages were in circulation in the United States and Utah region during data collection. The study ended on September 30, 2024 with results expected to be published in 2025.

**Conclusions:** Data from the BEEHIVE study will provide valuable real-world VE data for a heterologous dose of the Novavax COVID-19 Vaccine and homologous or heterologous dose of the Pfizer-BioNTech COVID-19 Vaccine after an mRNA-based COVID-19 primary series.

**Trial Registration:** Clinicaltrials.gov NCT06065176

## INTRODUCTION

### Background

Early in the COVID-19 pandemic, there was vigilant surveillance from regulatory authorities to monitor SARS-CoV-2 cases and the impact COVID-19 vaccines had on controlling viral spread. When public health emergency status was not renewed in May of 2023, surveillance mechanisms greatly diminished or stopped entirely [1]. With the continued evolution of immune-evasive variants, COVID-19 remains a public health concern [1]. Maintaining surveillance of vaccine effectiveness (VE) of updated COVID-19 vaccines targeting new strains [2] is informative for ongoing vaccination campaigns and the development of new vaccine formulations.

There are three COVID-19 vaccines approved for use by the United States (US) Food and Drug Administration (FDA) [3-5]. Two of these vaccines are mRNA-based (Pfizer-BioNTech COVID-19 Vaccine and Moderna COVID-19 Vaccine) and one is an adjuvanted recombinant spike protein (Novavax COVID-19 Vaccine). A key difference among these formulations is the use of mRNA encoding the SARS-CoV-2 spike protein [3,4] in the Pfizer and Moderna vaccines and the use of recombinant spike protein with Matrix-M™ adjuvant in the protein-based vaccine [5-7] in the Novavax vaccine. Prototype monovalent COVID-19 vaccines of each of these formulations targeting the ancestral Wuhan strain have gone through a series of updates (i.e., bivalent Wuhan and BA.4/5, monovalent XBB.1.5, and monovalent JN.1 lineage), since their initial authorization, to target circulating lineages of relevance at the time [8]. There are limited clinical and real-world data on the VE of XBB.1.5-based vaccines [9,10] and a better understanding is needed of the protection these formulations may provide.

### Objectives

The Booster Epidemiological Evaluation of Health, Illness and Vaccine Efficacy (BEEHIVE) randomized, controlled study (NCT06065176) was developed to assess VE of the 2023–2024 formulations (against XBB.1.5) for the Pfizer-BioNTech and Novavax COVID-19 vaccines in a real-world setting [11]. Study protocol design and endpoints are described here, as well as the novel methods of recruitment and ways to maintain participant engagement that were employed.

## METHODS

### Study Design

The BEEHIVE study was designed to enroll 1500 participants. Participants were aged ≥18 years, were from Salt Lake City, Utah (including a 60-mile radius of surrounding areas), had previously received ≥2 doses of an mRNA-based COVID-19 vaccine authorized by the US FDA, and qualified based on eligibility criteria described in **Textbox 1** and **Textbox 2**.

#### Textbox 1. Inclusion criteria.

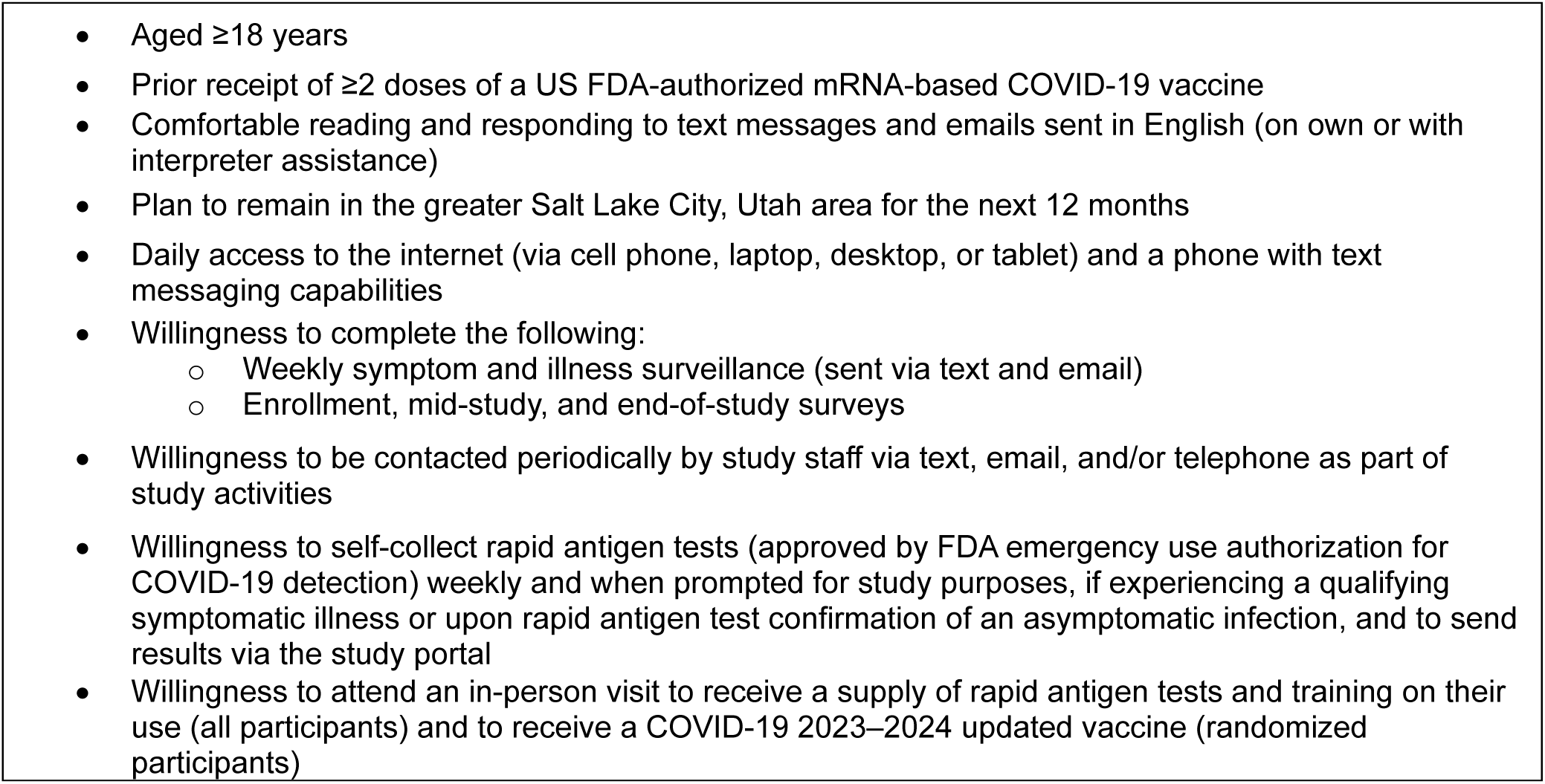

#### Textbox 2. Exclusion criteria.

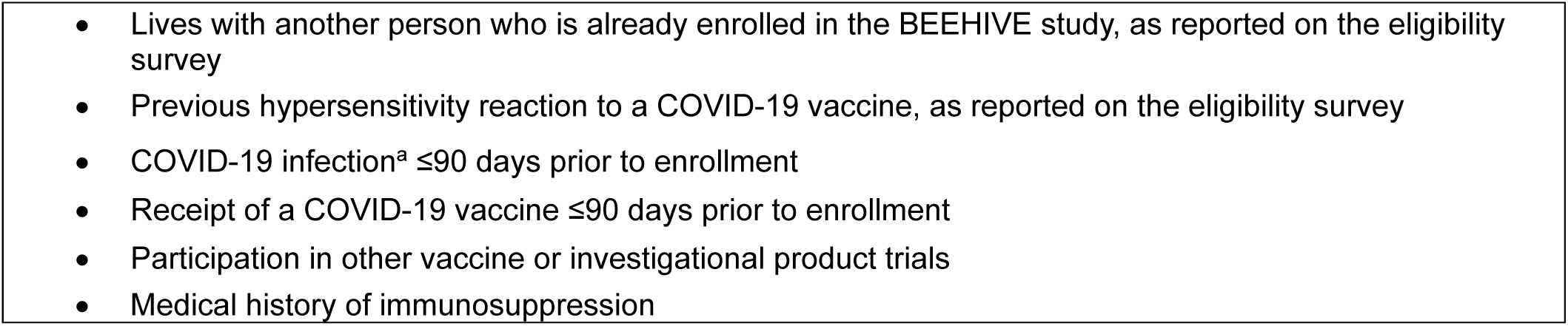
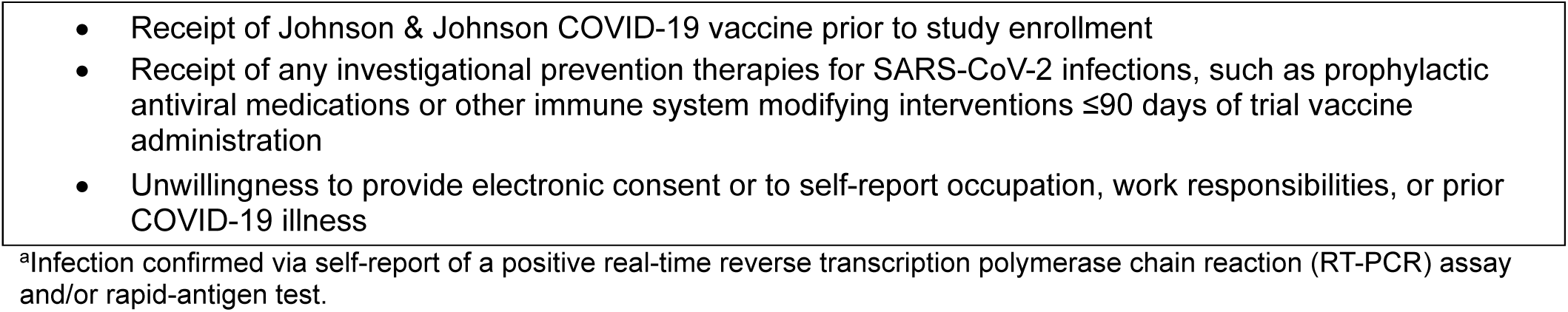

Study groups were comprised of two randomized, blinded groups who received the 2023–2024 formula of either the Novavax COVID-19 Vaccine or Pfizer-BioNTech COVID-19 Vaccine, or a non-randomized comparator group of volunteers who participated in the study, but did not elect to receive a study vaccine or any 2023–2024 COVID-19 vaccine formula during the study/surveillance period (**Figure 1A**).

**Figure 1.**
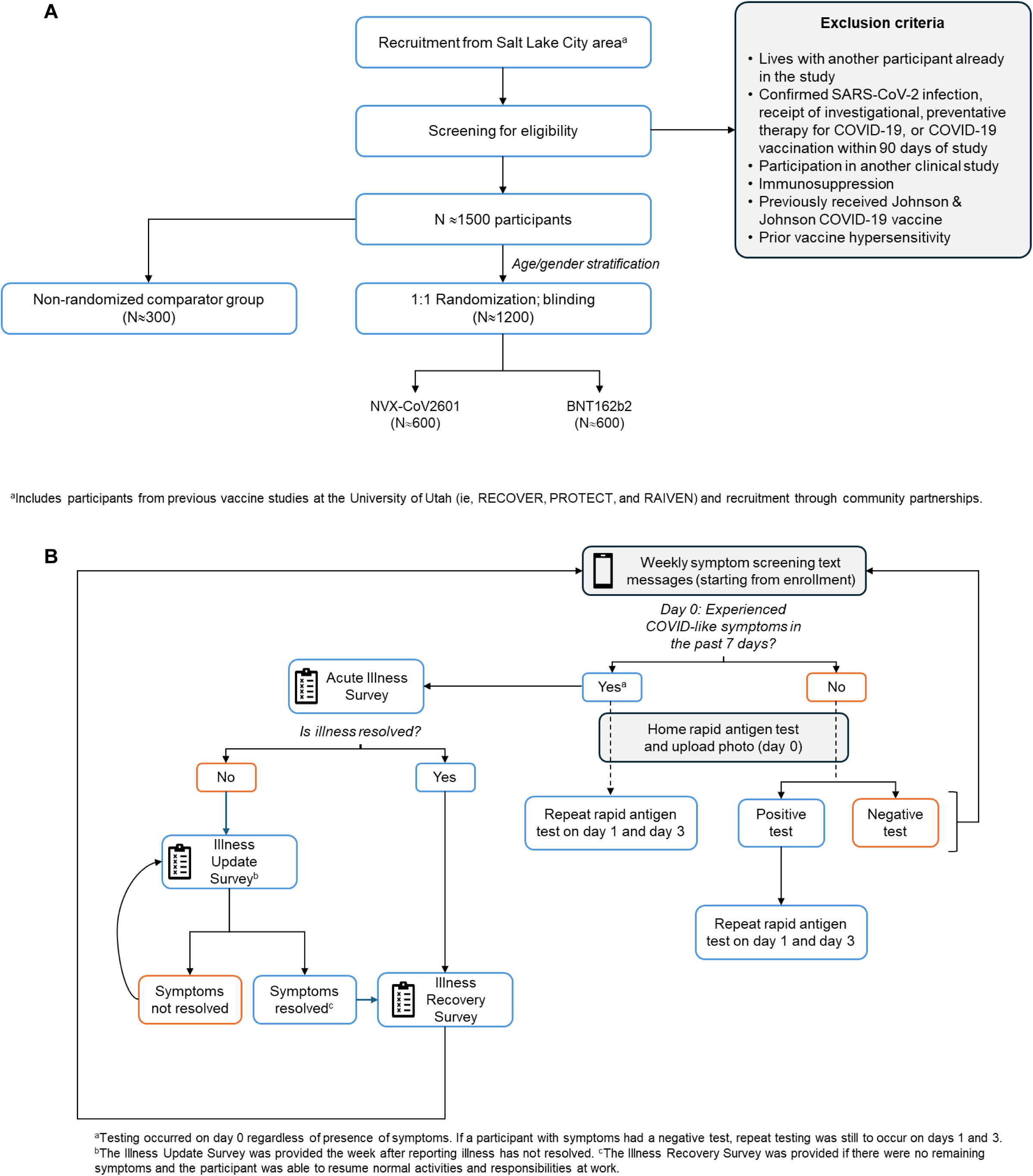
Study design. (A) Recruitment, screening, and randomization schema. (B) Decision tree for symptom and rapid antigen test monitoring. For this study, the interpretation of results is based on the participant’s status (eg, asymptomatic or has CLI symptoms) on the first day (day 0) of each testing. Additional tests on days 1 and 3 were required if a participant had symptoms on day 0, and if a test was positive on day 0, regardless of symptoms status. To be considered a true negative case, all three tests must result as negative.

### Participant Recruitment

Several unique mechanisms were used to facilitate recruitment. Radio and television advertisements were deployed in the Salt Lake City area and flyers for the BEEHIVE study included patient groups representative of populations that could match eligibility criteria (**Multimedia Appendix 1**). Participants from previous COVID-19 studies (i.e., RECOVER, PROTECT) [12,13] or an influenza vaccine study (i.e., RAIVEN) [14] who consented to be contacted for future studies were sent recruitment invitations. Outreach to support recruitment efforts included joining occupational health vaccination campaigns and other existing mass-vaccination outreach events, advertising through social media and local news outlets, having booths at community events, and deploying mobile vaccination stations. Collaborations were established with local community groups, clinics, businesses, universities, senior centers, and fire and police departments. Potential participants received a link to the online eligibility survey via email and were provided eligibility status upon survey completion. After completion of consent and Health Insurance Portability and Accountability Act (HIPAA) authorization forms, eligible participants were invited to generate an online portal account, complete the Enrollment Survey, and schedule an in-person enrollment visit online (**Multimedia Appendix 2**).

### Vaccination History

For participants who provided consent to having their vaccine records obtained by the study staff, the state immunization information system or registry was accessed to obtain COVID-19 and influenza vaccination status and history. Participants were also asked to provide their COVID-19 and influenza vaccination history through self-report and provide vaccine documentation to the study.

### Study Objectives

There were two aims of the BEEHIVE study. The primary aim was to compare VE between study-vaccinated participants with a comparator group who declined an updated COVID-19 vaccine (2023–2024 formula) in preventing symptomatic SARS-CoV-2 infection. The secondary aim was to determine the relative VE (rVE) of the 2023–2024 formula of the Pfizer-BioNTech mRNA and Novavax COVID-19 vaccines in preventing symptomatic SARS-CoV-2 infection.

Secondary objectives included the effect of the number of prior COVID-19 vaccinations on rVE of the Novavax COVID-19 Vaccine. Additional secondary objectives were to assess predictors of asymptomatic versus symptomatic SARS-CoV-2 infection, as well as clinical characteristics and outcomes associated with COVID-19, such as: illness duration and severity, socio-demographic and health characteristics associated with prolonged or severe illness; impact of COVID-19 on indicators of functioning (e.g., missed work, ability to complete normal work and home activities, and working while ill); the proportion of COVID-19 illnesses that were medically attended, and the factors associated with seeking medical care and treatment. Illness characteristics and duration for primary infection versus re-infection will be compared and whether VE is modified by socio-demographic characteristics, occupation, health status, or other risk factors. Goals of additional outcomes analysis are to learn if the 2023–2024 vaccine formula modifies illness severity and duration in those with breakthrough infection. Data related to the incidence of post-COVID conditions (PCC) were collected, along with identifying factors associated with symptoms of PCC, if these symptoms differed among randomized and comparator study groups, or if symptoms of PCC were modified by number of prior vaccine doses. Finally, the knowledge, attitudes, and practices (KAPs) of participants related to 2023–2024 vaccines are to be characterized and the associations between KAPs and subsequent vaccination behaviors (including vaccine refusal, hesitancy, or incomplete adherence to vaccination recommendations) will be examined.

### Ethical Oversight

Prior to study implementation, the protocol, informed consent form, participant education and recruitment materials, data collection instruments, and other documents associated with the protocol were approved by the University of Utah institutional review board. Electronic signatures were obtained for consent and HIPAA authorization forms, and participants were able to access consent forms in their portal accounts, as well as receiving an e-signed copy via email. All participants provided consent and were allowed to withdraw at any time. The research data platform and study database were maintained by Westat, Inc. (Rockville, Maryland).

### Randomization and Blinding

A computerized random-number generator produced the randomized list prior to study enrollment. Randomly permuted group blocks (designated A–F) of size 6 with 3 of each vaccine type in each block were generated for 1:1 assignment. Randomization was stratified by sex and age (18–49 years and ≥50 years). Eligible participants who provided consent indicating agreement to receive an updated 2023–2024 vaccine were able to schedule their first study visit and vaccination appointment. Participants were randomized to either the Novavax COVID-19 Vaccine group or the Pfizer-BioNTech COVID-19 Vaccine group; however, based on appointment selection and attendance, some participants who consented may not have completed randomization or received a study vaccine. Enrolled participants who provided consent to the study but declined receipt of the 2023–2024 formulation were included in the non-randomized comparator group, until enrollment was full. Participants who changed their minds at the time of their first study visit were able to switch groups (i.e., initially agreed to be randomized to receive a vaccine, but then switched to the non-randomized comparator group, and vice versa) at the time of their appointment.

Randomized participants, study investigators, and study personnel were blinded to the type of vaccine received during the study and multiple procedures were performed to ensure maintenance of this blinding. Specifically, vaccine packaging, administration, and documentation practices were standardized to avoid any distinguishable characteristics between the two vaccines. Additionally, all study staff, except licensed healthcare providers who administered the vaccines, were blinded and received training about the importance of consistent data collection and blinding. Vaccines were prepared in a designated space obscured from participants and study personnel. Study personnel who administered the vaccines were unblinded and underwent specialized training to ensure vaccine type was not inadvertently disclosed; these personnel were not involved in any data collection or analysis and only interacted with the participant to confirm their identity and administer the vaccine. The Westat randomization coordinator and limited data management staff were also unblinded to facilitate development and maintenance of the randomization list and systems. These staff were not involved in data collection or analysis, or interaction with participants. Participants and study investigators were unblinded at the end of data collection and the end of the study, respectively. All study-related documents and samples contain a unique identifier for each person to preserve blinding.

### Interventions

Randomized participants received a single dose of study vaccine (0.3 ml for Pfizer-BioNTech or 0.5 ml for Novavax) administered in the deltoid and were to be monitored for 15 minutes after vaccination for any allergic reactions. An investigational new drug application was submitted and approved by the US FDA for use of the Novavax COVID-19 Vaccine in this study since it was under an Emergency Use Authorization at the time of the study.[5]

### Surveillance: Surveys

In the Enrollment Survey, the participants self-reported on their socio-demographic characteristics, health status and behaviors, occupational status and employment history, chronic medical conditions, COVID-19 and influenza vaccination history, and previous SARS-CoV-2 infections.

A post-vaccination survey was shared 24- and 48-hours and 6 days after study vaccination to obtain information on potential vaccine side effects. Active surveillance of outcomes occurred for participants on the same day each week, beginning the week following the enrollment/study vaccination visit and occurred for 24 weeks (**Figure 1B**).

Symptoms were assessed weekly through Surveillance and Illness Messaging Surveys through 24 weeks after enrollment/study vaccination. Surveillance included weekly subjective assessment of COVID-19–like illness (CLI), which was defined as experiencing at least one of the following symptoms: chills, malaise, fatigue, headache, cough, shortness of breath, sore throat, runny nose/nasal congestion, nausea/vomiting, diarrhea, muscle/body aches, change in smell or taste.

Follow-up surveys were deployed at designated timepoints and allowed participants to provide socio-demographic and illness updates, assess KAPs, and collect information on PCC (**Textbox 3**).

#### Textbox 3. Topic areas of follow-up surveys

|  |
| --- |
| • Report symptoms related to PCC (e.g., lasted longer than 1 month), if applicable |

During the surveillance period, if a participant indicated a CLI symptom on a weekly survey or reported an acute illness between their designated weekly surveillance contacts, they were directed to the Acute Illness Survey (**Figure 1B**). In this survey, they were asked to identify qualifying symptoms from a detailed checklist, confirm the date of onset, and indicate whether the symptoms were ongoing. If the participant did not confirm symptoms of a CLI, they returned to weekly surveillance. If the symptoms were ongoing, participants were prompted to collect a specimen and submit the results according to study procedures. Illness Update and Illness Recovery Surveys were deployed to monitor symptoms until a participant reported ≥90% recovery (**Figure 1B**). Survey data were collected electronically through a text messaging interface and online surveys (**Multimedia Appendix 2**).

### Surveillance: Testing

All participants received OHC COVID-19 Antigen Self-Test kits (Osang Healthcare, South Korea) to use every week with instructions for how to use the kit, interpret results, and how to upload results to the study portal (**Multimedia Appendix 1**). Anterior nasal swab specimens were to be self-collected weekly on the same day (day 0) and a photo of results uploaded, regardless of symptoms (**Figure 1B**). If the participant reported an acute illness and/or collected an illness specimen on or outside their assigned routine specimen day, they were asked to collect a 2^nd^ specimen on the next day (day 1) and a 3^rd^ specimen the two days after that (day 3). For this study, the interpretation of test results was based on the participant’s symptom status the first day of testing (day 0) and whether they did or did not have CLI symptoms.

### Safety Assessments

Safety assessments included monitoring and recording of solicited (local and systemic reactogenicity events) and unsolicited adverse events (AEs), serious AEs (SAEs), and AEs of special interest (AESIs). Solicited AEs were collected through the Post-Vaccination Survey. Unsolicited AEs could be reported any time from study vaccination to the participants’ completion of the last study-related procedure. Relationship to study vaccine (related or not related), whether the event was expected or unexpected, whether the event was serious or not, and intensity of AEs and SAEs was investigator-determined; severity was categorized as mild (grade 1), moderate (grade 2), severe (grade 3), life-threatening (grade 4), or death (grade 5). Protocol-defined AESIs were myocarditis and pericarditis and were collected by participant report only; participants received an informational sheet describing these symptoms. Safety reporting related to the investigational new drug (IND) status of the Novavax COVID-19 Vaccine included sharing Suspected Unexpected Serious Adverse Reactions and relevant medication errors to the appropriate regulatory authorities.

### Statistical Analyses

The primary population for VE analyses will be the modified intent-to-treat (mITT) population, which includes all randomized participants who received a study vaccine and responded to ≥1 active surveillance survey, and all participants in the comparator group. Any participant with a CLI-associated SARS-CoV-2 infection confirmed by COVID-19 rapid antigen test 0–6 days after vaccination will be excluded from the mITT population. Analyses with the per-protocol population will also be performed. This population includes participants who met eligibility criteria, received a study vaccine per protocol, participated in study surveillance by responding to at least one surveillance contact during the SARS-CoV-2 circulation period (active surveillance), and did not receive another SARS-CoV-2 vaccine, outside of the study, during the study period. If participants were in the non-vaccinated comparator group but subsequently received a 2023–2024 updated vaccine, they will be censored at that point.

With 1500 participants enrolled (1200 randomized and 300 non-randomized comparator), assuming ≤15% attrition, there would have been 80% power to detect a VE of ≥49.4% for the primary aim (ie, VE between randomized and comparator participant groups), examined with a 2-sided alpha=0.05 superiority test. The calculation was based on an estimated incidence rate of 80 per 100,000 person-days for the comparator group. For the secondary aim, noninferiority of the Novavax COVID-19 Vaccine versus the Pfizer-BioNTech COVID-19 Vaccine will be declared if the lower one-sided 95% confidence limit for rVE is above −50%. With 600 participants per vaccine group and an assumed infection rate of 60/100,000 person-days in the Pfizer-BioNTech COVID-19 Vaccine arm and a rVE of 10% for the Novavax COVID-19 Vaccine versus the Pfizer-BioNTech COVID-19 Vaccine (ie, Novavax > Pfizer-BioNTech)[15,16], there would be 80% power for the non-inferiority objective. The positive rVE considered in the sample size calculation is based on the rVE estimated in large retrospective studies[15,16] showing the Novavax COVID-19 Vaccine offering added protection against medically-attended and severe SARS-CoV-2 infection. Sample size calculations for study aims were performed in PASS 2023 with Cox Proportional Hazards Model used for HRs. Certain secondary objectives will be underpowered if there are not a sufficient number of non-symptomatic cases, reinfections, and/or PCC during the study period.

Participant demographics and characteristics will be summarized descriptively in the mITT population. The Cox proportional hazards model will be used to analyze time-to-event data to evaluate VE in preventing symptomatic COVID-19 infection. The observation period started 7 days after the receipt of the updated 2023–2024 vaccine for the randomized groups and at the enrollment date for the comparator group. The event onset time is defined as the earlier of the date of a positive rapid antigen test or the date of CLI symptom onset reported within a week of the positive rapid antigen test. Covariates of age and vaccination dose history will be included in the model to control for confounding. All analyses will be performed using SAS Software^®^ version 9.4 or higher, or R version 4.1.2 or higher.

### Remuneration and Engagement

Participants received compensation in the form of Amazon, Inc. (Seattle, Washington) gift cards with the value being relative to the number of study activities completed (**Multimedia Appendix 1**). Newsletters were shared throughout the study to investigators and participants and included information such as enrollment numbers, how to remain compliant/up-to-date with study protocol and surveys, and updates on COVID-19 activity in the area. Study branding was consistently presented from recruitment and throughout the study.

## RESULTS

### Participant Enrollment and Data Collection

Participant screening occurred from November 17, 2023, through March 14, 2024. Monitoring occurred from enrollment through study end on September 30, 2024. A total of 1188 participants were enrolled, 909 in the vaccine group (Novavax COVID-19 Vaccine, n=452; Pfizer-BioNTech COVID-19 Vaccine, n=457) and 279 in the comparator group. Data analyses are ongoing, and results are expected to be published in 2025.

### SARS-CoV-2 Circulation

Data collection during the study occurred when XBB-, JN.1-, KP.2-, and KP.3-lineage SARS-CoV-2 variants were circulating in the US and Utah region (**Figure 2**).[17]

**Figure 2.**
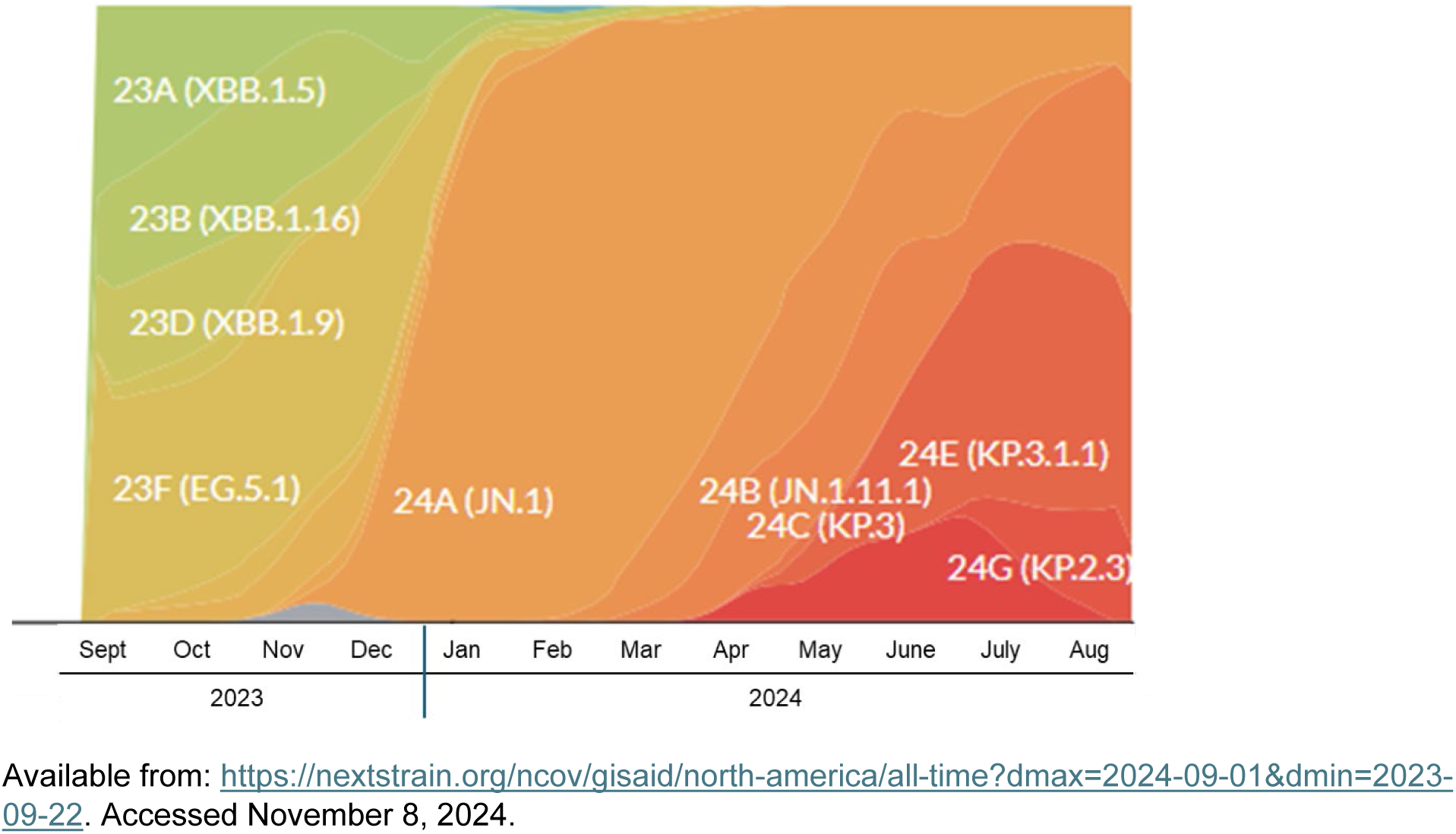
Circulating SARS-CoV-2 strains during the BEEHIVE study.

## DISCUSSION

The BEEHIVE study will provide real-world VE data for the Novavax COVID-19 Vaccine as an additional dose after at least a primary series of an mRNA-based COVID-19 vaccine and data on the comparison with the Pfizer-BioNTech COVID-19 mRNA COVID-19 Vaccine. Outcomes may provide information on the benefits of heterologous dosing or using a mix of different vaccine formulations for administration beyond primary vaccination.

Access to participant cohorts from previous studies allowed recruitment of individuals who were more likely to participate, as well as facilitated access to socio-demographic information and vaccination history. Mobile vaccine clinics, visiting community events, and collaborations with civic groups assisted in engaging typically underrepresented communities (eg, elderly, communities of color)[18,19] than standard clinical trial site strategies. Electronic surveillance and the use of online/text surveys facilitated data collection and maintenance of participant engagement throughout the 24-week surveillance period.

A potential limitation of this study includes a lack of participants in upper age categories. Additionally, the variability in SARS-CoV-2 circulation and introduction of variant strains during the study period would impact infection rates and symptom severity. Notably, the testing kits, which were identical for all study participants, had received emergency use authorization from the US FDA for Omicron and subvariant testing. The single-site nature of the trial may restrict geographic diversity among the participants; however, feasibility of a real-world study design was established and can be adapted for potential future, multi-site studies. Since rapid-antigen tests were utilized given the real-world design of the study, repeat testing procedures were included in the design to help mitigate concerns over not using a more robust clinical assay (eg, polymerase chain reaction). Additionally, assuming superior rVE of NVX compared to mRNA vaccines based on study results of medically attended COVID-19 infection may not directly translate into potential benefits protecting from symptomatic, SARS-CoV-2 infection; therefore, future non-inferiority studies should consider sample size calculations using a rVE of 0 to ensure a more robust study.

Continued surveillance of COVID-19 cases and VE data for vaccine formulations updated based on predominant SARS-CoV-2 strains are a necessary to inform public health policy and management. The BEEHIVE study will provide valuable real-world VE data for a population representative of those who have received an updated COVID-19 vaccine, as well as information on receipt of a heterologous dose of the Novavax COVID-19 Vaccine after receipt of ≥2 mRNA-based COVID-19 vaccines.

## Acknowledgements

We thank each of the study participants who volunteered for this study. We thank Chengbin Wang, MD, PhD, and Mark Thompson, PhD, for their initial study support. This study was funded by Novavax, Inc. Medical writing and editorial support were provided by Kelly M. Fahrbach, PhD, CMPP, and Ebenezer M. Awuah-Yeboah, BS, of Ashfield MedComms (New York, USA), an Inizio company. Additionally, editorial support was provided by Anar Murphy, PhD, CMPP, of Novavax, Inc.

## Data Availability

Study information is available online at https://www.clinicaltrials.gov/study/NCT06065176.

## Author Contributions

**Conceptualization** – SKY, SB-W, ER, SWB, MST, ALP, GLE, ASS

**Data curation** – HZ, EAKR, SB-W, JM, RC, JW

**Formal analysis** – SKY, SB-W, EAKR, RVF, SWB, HZ

**Funding acquisition** – BW, MDR, SKY, MST, ALP, SWB

**Investigation** – SKY, GLE, ALP, MST, JM, RC, JW, JG

**Methodology** – SKY, SB-W, EAKR, SWB, MST, ALP, GLE, ASS, HZ, JM, JG

**Project administration** – SKY, SB-W, RVF, SWB, MST, ALP, GLE, RC, JW, JM, JG, ASS

**Resources** – BW, SKY, MST, ALP, GLE, ASS, SWB

**Software** –SKY, ALP, GLE, JM, RC, JW, SB-W, SWB, RVF, EAKR, HZ

**Supervision** – SKY, MDR, JM, JG, SWB, SB-W, RVF, ASS

**Validation** – HZ, EAKR, SB-W, SKY, MDR

All authors reviewed and edited drafts and approved the final draft for submission.

## Abbreviations

AE: adverse event
AESI: adverse event of special interest
BEEHIVE: Booster Epidemiological Evaluation of Health, Illness and Vaccine Efficacy
CLI: COVID-like illness
FDA: Food and Drug Administration
HIPAA: Health Insurance Portability and Accountability Act
IND: investigational new drug
KAP: knowledge, attitudes, and practices
mITT: modified intent-to-treat
PCC: post-COVID conditions
PCR: polymerase chain reaction
SAE: serious adverse event
VE: vaccine effectiveness
US: United States

**Multimedia Appendix 1**: Recruitment and engagement materials

Recruitment Ad; Recruitment Flyer; Renumeration guidelines; Test and Upload Instructions; Interpretation of test results

**Multimedia Appendix 2**: Electronic interfaces Examples of account and survey home pages

