## Appendix 1 for "Real-world effectiveness and non-inferiority evaluation and comparison of mRNA- and protein-based COVID-19 vaccines: a randomized study protocol (BEEHIVE)"

### RECRUITMENT ADVERTISEMENTS

#### TELEVISION AD

**U OF U HEALTH| 15 seconds | COVID BOOSTER STUDY**

| AUDIO | VIDEO |
|---|---|
| The University of Utah invites residents of the greater Salt Lake City area to join our COVID-19 booster study whether or not you plan to get a booster.<br><br>If you received at least two doses of COVID-19 vaccine in the past, you may qualify.<br><br>You could receive a free COVID-19 booster if you would like one. You can be in this study even if you don't want a booster.<br><br>You will receive free at-home COVID-19 test kits for participating in the booster study. You could receive up to $550 for completing study activities.<br><br>Email or call right now to learn more! | [Graphics on right side of screen for duration of the spot with footage of teachers, food workers, bartenders, retail workers, construction workers, paramedic, and older adults.]<br><br>[U of U Health logo]<br><br>COVID-19 BOOSTER Study<br><br>Adults 18 years and older who received at least two doses of COVID-19 vaccine in the past may qualify<br><br>Chance to receive up to $550 for completing study activities<br><br>EMAIL \|<br><br>CALL \| (801) 203-0320<br><br>[Study logo] |

**Pure Flyer Video. Voiceover explaining the study (BeehiveFlyerVid.mp4)**

Voiceover: Hi, are you interested in being part of an important research study? The BEEHIVE study at the University of Utah is comparing how well two different FDA-authorized booster vaccines protect people from COVID-19 and whether getting a booster provides better protection against COVID-19 than not getting a booster. You'll have the opportunity to get a COVID booster as part of this study, or you can join the study and not get boosted. If you decide to join the BEEHIVE study, you'll be asked to complete activities such as weekly COVID tests and surveys about your health. You'll also have the chance to receive up to $550 in Amazon gift cards. For more information and to see if you qualify, visit the study website at www.BEEHIVEstudy.com.

### BEEHIVE

Booster Epidemiological Evaluation of Health, Illness and Vaccine Efficacy

We are looking for adult volunteers aged 18 years and older living in the greater Salt Lake City area to participate in a research study that will compare how well two different FDA-authorized boosters protect against COVID-19.

**Enrollment begins Fall 2023**

#### What is involved?

- Receive up to $550
- You will choose whether or not to receive an FDA-authorized COVID-19 booster
- Responding to online surveys each week and when you are sick
- Free at-home COVID-19 rapid testing and self-reporting
- Answering a survey at the beginning, middle and end of the study

**Contact us to learn more and see if you qualify:**


(801) 203-0320

#### We need your support!

HEALTH
UNIVERSITY OF UTAH

**Table**. Renumeration guidelines

| Study activity | Amazon gift card value (USD) |
|---|---|
| Enrollment survey | $30 |
| Completed enrollment visit | $50 |
| Post-vaccination survey | Days 1 and 2: $12<br>Day 7: $20 |
| Post enrollment weekly symptom and illness surveys and COVID-19 rapid antigen test photo submission | $14 (total up to $336 for 24 weeks) |
| Mid-study and End-of-study surveys | $30 (each) |
| Long-COVID surveys (if applicable) | $15 (each, up to $30) |
| **TOTAL** | **Up to $550** |

BEEHIVE
Booster Epidemiological Evaluation of Health, Illness and Vaccine Efficacy

HEALTH
UNIVERSITY OF UTAH

### Instructions for Your At-Home COVID-19 Test & How to Upload Results

**1** Please confirm that your test kit contains:

- a white packet
- a silver packet
- a nasal swab

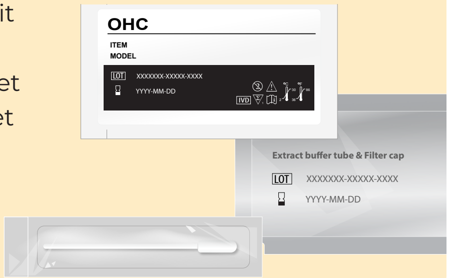


**2** Blow your nose. Sanitize or wash and dry your hands thoroughly.

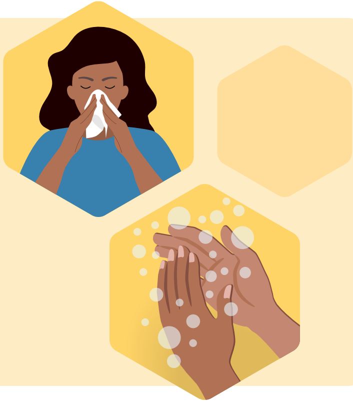

**3** Open the silver packet.

Then, carefully peel off the foil seal from the Extraction Buffer Tube. Keep it upright so it doesn't spill. If any liquid spills, discontinue and open a new test.

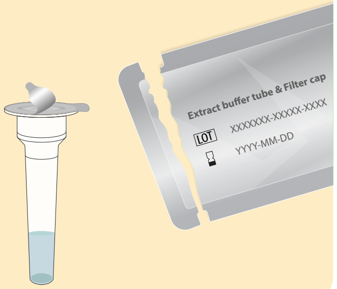


**4** Remove the sponge swab from the wrapper by the handle. Be careful not to touch the tip.

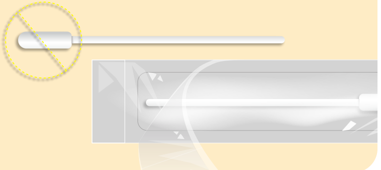

**5** Insert the entire soft end of the swab approximately ½-inch to ¾-inch into your nostril.

Rotate at least **5 times** for **15 seconds**. If you encounter resistance, do not insert any further. Make sure you swab each inside wall of the nostril. Do not just spin the swab.

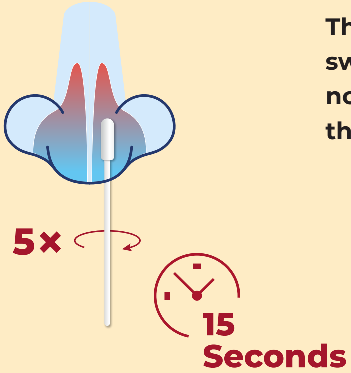


**Then, insert the same swab into your other nostril and repeat these same steps**.

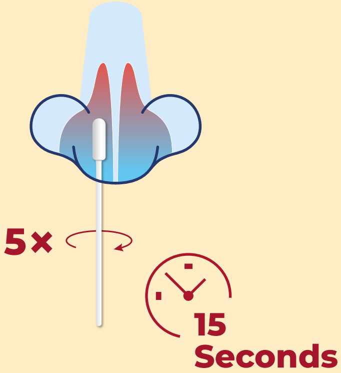


**6** Place the swab into the tube with the swab tip facing downward.

Stir the swab around in the liquid at least **10 times**.

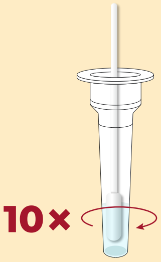


**7** As you remove the swab, pinch the sides of the tube with your other hand to squeeze the remaining liquid from the swab. Discard the swab. *Failure to squeeze the tube can lead to incorrect results due to excess buffer left in the swab tip.*

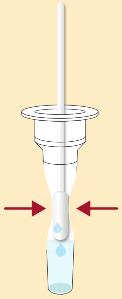

**8** Place the filter cap on the tube with the long nozzle sticking out of the top of the tube. Push until the cap is flush with the tube and no liquid can leak out.

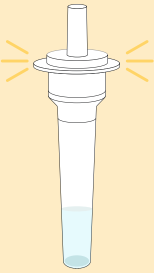

**9** Open the white packet and place the test cassette on a flat surface. Now, hold the sample tube upright above the test cassette and squeeze exactly **4 drops** into the oval-shaped sample well. *Adding more than 4 drops may result in incorrect results.*

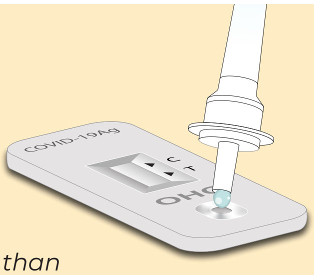


#### Wait 15 Minutes

**10** Wait exactly **15 minutes**. Do not touch or move your test during this time. This is a good time to fill out your weekly survey on the study portal.

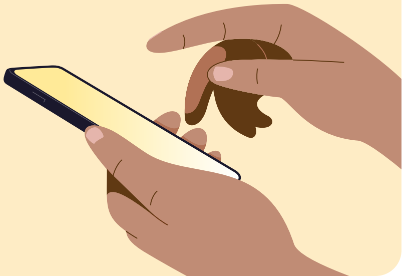

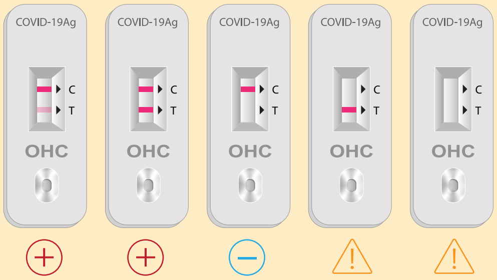


**11** After 15 minutes, look at your test cassette.

The **C** on your test stands for **Control**. If there is a line next to the C, the test is valid. If there is no line, the test is invalid and you will need to re-test.

The **T** on your test stands for **Test**. If there is a line next to the T, the test is positive for COVID-19. If there is no line next to the T, the test is negative for COVID-19.

*Inaccurate test results may occur if results are read before 15 minutes or after 20 minutes.*

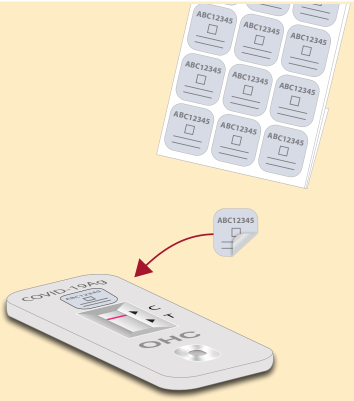


**12** Use one of the stickers you were given to label your test in the top space on the test cassette. The stickers do not have to be used in a certain order.

**13** Take a photo of your test result. Make sure the test cassette and sticker are clearly visible in the photo.

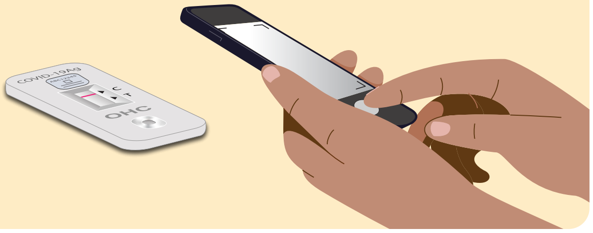

**14** Upload this photo to the secure BEEHIVE portal and input your test results and the sticker number you used.

Discard all test materials.

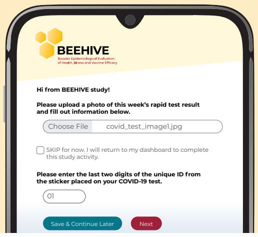
