## Appendix 2 for "Real-world effectiveness and non-inferiority evaluation and comparison of mRNA- and protein-based COVID-19 vaccines: a randomized study protocol (BEEHIVE)"

### Eligibility Survey Home Page

Log in

BEEHIVE

Booster Epidemiological Evaluation of Health, Illness and Vaccine Efficacy

Thank you for your interest in the Booster Epidemiological Evaluation of Health, Illness and Vaccine Efficacy (BEEHIVE) Study! We are conducting a research study beginning in Fall 2023. The purpose of this study is to compare how well two different COVID-19 boosters work to protect people 18 years of age and older against COVID-19. Both boosters have received Food and Drug Administration (FDA) Emergency Use Authorization* for adults aged 18 years and older in the United States.

To find out if you are eligible to participate in this study, click See If I'm Eligible to answer a few questions. If you would like to learn more about this study, select Frequently Asked Questions.

See if I'm Eligible | Frequently Asked Questions

#### What's involved

BEEHIVE
Booster Epidemiological Evaluation of Health, Illness and Vaccine Efficacy

BEEHIVE
Booster Epidemiological Evaluation of Health, Illness and Vaccine Efficacy

HEALTH
UNIVERSITY OF UTAH

### Consent and HIPAA Authorization Form

#### Adult Electronic Consent

**Research Study Title:** Booster Epidemiological Evaluation of Health, Illness and Vaccine Efficacy (BEEHIVE) Study: Randomized Trial to Compare the Clinical Efficacy of Novavax vs. mRNA COVID-19 boosters among adults 18-49 and 50+ years in the United States

**Study No.:** [redacted]

**Principal Investigator:** Sarang Yoon, DO

**Address:** 250 East 200 South, Suite 100 Salt Lake City, UT 84111

This consent and HIPAA authorization form will give you information to help you decide if you want to take part in an important research study.

##### Key Information:

The first section of this document contains key points that the research team thought you would find important. The study is described in more detail after this section. If you have any questions, please call (385) 274-7354 to speak to study staff.

We are inviting you to participate in a research study that will compare how well COVID-19 booster vaccines protect people from the SARS-

### Account Creation Home Page

BEEHIVE
Booster Epidemiological Evaluation of Health, Illness and Vaccine Efficacy

#### Create your account below

 (this is your username)

Password*

Confirm Password*

Passwords must be at least 8 characters.
Passwords must have at least one non alphanumeric character ('&', '$', '!').
Passwords must have at least one digit ('0'-'9').
Passwords must have at least one lowercase letter ('a'-'z').
Passwords must have at least one uppercase letter ('A'-'Z').

Submit

### Enrollment Survey Home Page

**BEEHIVE**
Booster Epidemiological Evaluation
of Health, Illness and Vaccine Efficacy

#### INSTRUCTIONS:

**Thank you for agreeing to participate in the BEEHIVE study. You have logged in to the "Enrollment Survey". This survey will take about 20 minutes or less to complete. To navigate the survey, please use the Next and Previous buttons at the bottom of the page (do not use the browser Next/Back buttons).**

Save & Continue Later

Next

For questions about your rights and welfare as a research participant or for a research-related injury, please call the Westat Human Subjects Protections Office at 801-203-0320. Please leave a message with your first name, the name of the research study (BEEHIVE) that you are calling about, and a phone number beginning with the area code. Someone will return your call as soon as possible.

novavax
